# Lack of association between angiotensin converting enzyme inhibitors and angiotensin receptor blockers and pain improvement in patients with oral cancer

**DOI:** 10.1101/2020.05.05.20091868

**Authors:** K.N. Du, A.J. Shepherd, I.V. Ma, C.J. Roldan, M. Amit, L. Feng, S. Desai, Juan P. Cata

**Affiliations:** Department of Internal Medicine. Baylor College of Medicine– Houston, Texas. USA; Department of Symptom Research. The University of Texas MD Anderson Cancer Center – Houston, Texas. USA; University of Nevada, Reno School of Medicine; Department of Pain Medicine. The University of Texas MD Anderson Cancer Center – Houston, Texas. USA; Department of Head and Neck Surgery. The University of Texas MD Anderson Cancer Center – Houston, Texas. USA; Department of Biostatistics. The University of Texas MD Anderson Cancer Center – Houston, Texas. USA; Department of Anesthesiology and Pain Medicine. The University of Texas MD Anderson Cancer Center – Houston, Texas. USA; Anesthesiology and Surgical Oncology Research Group. Houston, Texas. USA

**Keywords:** oral cancer, pain, angiotensin receptor blockers

## Abstract

**Background:** There is a growing body of literature implicating angiotensin II in the modulation of tumor associated inflammation and pain. However, the impact of angiotensin converting enzyme inhibitors (ACEis) and angiotensin II receptor blockers (ARBs) on this pathway has not yet been studied in oral cancers. Our objective is to investigate the role of ACEi and ARB pharmacotherapy on preoperative pain and inflammatory biomarkers, neutrophil to lymphocyte ratio (NLR) and monocyte to lymphocyte ratio (MLR), in patients with oral cancer.

**Methods:** We performed a retrospective study on patients who underwent oral cancer surgery. Wilcoxon rank sum test or Kruskal-Wallis analysis were used to evaluate differences in demographic, tumor-related, and preoperative characteristics and among patients using ARBs, ACEis, and no treatment. Multivariable analysis was fitted to estimate the effects of important covariates on severe preoperative pain.

**Results:** 162 patients with oral malignancies were included in the study. After adjusting for significant covariates, patients with perineural invasion were found to have higher levels of pain (p = 0.0278). Likewise, patients taking ARBs were found to have lower levels of perineural invasion (p = 0.035). Our analysis did not demonstrate a significant difference in pain levels when comparing ARBs or ACEis to the no treatment group (p= 0.250). Furthermore, ARB or ACEi use did not significantly alter preoperative NLR (p = 0.701) or MLR (p = 0.869).

**Conclusions:** When compared to no treatment, ARBs and ACEis are not associated with significant analgesic effect or decreased inflammatory scores (NLR, MLR).

## Introduction

Oral and lip cancers rank among the 15^th^ most common malignancies worldwide.[1] Approximately 550,050 patients in the world are diagnosed with oral cancers each year.[1] Oral cancers are more common among men, older age groups and those with cofactors such as tobacco and alcohol use.[2] This patient population is also at risk for cardiovascular comorbidities. Therefore, angiotensin II receptor blockers (ARBs i.e, losartan, valsartan and omelsartan) and angiotensin converting enzyme inhibitors (ACEis, i.e., enalapril and linsinopril) are commonly prescribed to patients with oral cancers to treat hypertension, heart failure or prevent hypertensive nephropathy.[3]

Previous studies indicate that ACEis and ARBs modulate important signaling mechanisms involved in inflammatory and neuropathic pain.[4–7] Pain is one of the most common complaints of patients with oral malignancies and can be present even before any cancer treatment has been initiated.[8] The severity of pain in patients with oral cancer is rated as more intense than pain produced by other malignancies as it interferes with essential body functions including eating, talking, and swallowing, and can be refractory to conventional treatments.[9]

Multiple mechanisms appear to mediate pain in oral cancer including inflammation and nerve invasion. Preclinical studies by Scheff et al. indicate that tumor necrosis factor alpha (TNF-α), a known inflammatory cytokine, significantly contributes to tongue allodynia in mice injected with supernatant media from oral cancer cells.[10] TNF-α also upregulates chemokine ligand 2 (CCL2), a monocyte chemoattractant implicated in the potentiation of perineural invasion.[11, 12]. Bakst et al demonstrated that CCL2 secreted by the nerves recruits inflammatory monocytes to differentiate into macrophages and propagate perineural invasion in mouse cancer models.[12]

Additionally, other nociceptive mediators such as nerve growth factor (NGF) also promote nociception in mice bearing oral cancers. Ye et al. demonstrated in mouse models that NGF is implicated in changes in the expression of the transient vanilloid receptor 1 (TRPV1) in trigeminal ganglion cells, suggesting an interaction between NGF and TRPV1.[13] The expression of NGF in the neurons is regulated by the angiotensin II (AT2) receptor which in turn can modulate the function of TRPV1.[6] Moreover, the antagonism of the AT2 receptor also decreases the infiltration of CD3+ T cells and macrophages in the dorsal root ganglion (DRG), which correlates with a reduction in allodynia in animals with neuropathic pain.[6, 14, 15] Therefore, it appears that the AT2/NGF/TRPV1 interaction is an important mechanism in mediating nociception and inflammation. Currently, the possible impact of ACEis and ARBs drugs on pain associated to oral cancer has not been established.

The study was designed to evaluate a possible modulation role of ARBs and ACEis in pain associated to oral cancers. Due to the neuropathic and the inflammatory components of pain associated with this pathology, we hypothesized that patients actively taking ARBs and ACEis would present with lower pain intensity before surgery. Secondly, we also investigated whether the use of ARBs or ACEis had any impact on inflammatory markers. Since AT2 is a known inflammatory mediator via CD3+ T cells and macrophages, we hypothesized that patients taking ARBs and ACEis would present with lower levels of inflammation and decreased leukocyte proliferation, which we assessed via the neutrophil-to-lymphocyte ratio (NLR) and monocyte-to-lymphocyte ratio (MLR).

## Methods

After approval from the University of Texas MD Anderson Cancer Center Institutional Review Board (#PA16-1033), we performed a retrospective study that included a cohort of patients with oral cancers who underwent surgical treatment between January 2004 to January 2018. Patients 18 years of age or older were included, whereas patients with positive HPV status, and those with missing information regarding demographics, tumor pathology, preoperative pain, preoperative pharmacotherapy, and living status were excluded.

Information collected from the electronic medical records included age, gender, body mass index (BMI), American Society of Anesthesiologist’s (ASA) physical classification, stage of disease, history of smoking, human papillomavirus (HPV) status, tumor location, presence of perineural invasion, neoadjuvant chemotherapy, preoperative analgesics, and preoperative blood pressure pharmacotherapy (ACEi, ARBs, no treatment). Dependent variables recorded included self-reported preoperative pain intensity using a verbal numeric rating scale (VNRS; 0 - 3 = mild pain, 4-6 = moderate pain, 7-10 = severe pain) at the time of anesthesia assessment, preoperative neutrophil to lymphocyte ratio (NLR) and preoperative monocyte to lymphocyte ratio (MLR).

### Outcomes

In analyzing the effects of ARBs and ACEis on inflammation and nociception, the primary outcomes of our study were (1) preoperative oral cancer pain score as rated by the VNRS, and (2) level of inflammation as indicated by biomarkers, NLR and MLR. Preoperative NLR was defined as the ratio between the absolute neutrophil count and absolute lymphocyte count of routine blood samples drawn within fourteen days prior to date of surgery, closest to time of surgery. Preoperative MLR was defined as the ratio between the absolute monocyte count and absolute lymphocyte count of blood samples drawn within fourteen days prior to date of surgery, closest of time of surgery.

### Statistical Analysis

Summary statistics including mean, standard deviation, median, and range for continuous variables such as age, BMI, preoperative hemoglobin, NLR and MLR, and frequency counts and percentages for categorical variables such as ASA, T stage, N stage, severe preoperative oral cancer pain and recurrence/death status are provided. The Chi-square test or Fisher’s exact test was used to evaluate the association between two categorical variables. Wilcoxon rank sum test or Kruskal-Wallis was used to evaluate the difference in a continuous variable between/among patient groups. Multivariable logistic regression model was fitted to estimate the effects of important covariates on severe preoperative pain. A p-value < 0.05 was considered statistically significant. Statistical software SAS 9.4 (SAS, Cary, NC) and S-Plus 8.2 (TIBCO Software Inc., Palo Alto, CA) were used for all the analyses.

## Results

Of the 589 patients who underwent surgical resection for oral cancer, 162 patients met inclusion criteria. Demographic and clinical characteristics of patients based on their use of ARBs, ACEis, or neither pharmacotherapy (none) are summarized in Table 1. The median age was 60 years (range: 18~92), and the median BMI was 26.2 (range 14.1 ~ 46.3). There were 124 males, and the majority had a history of cigarette smoking (97.1%), and an ASA physical status of 3 (85.1%) (Table 1). The most common primary tumor location was the tongue (56.5%). Other oral cancer locations included the floor of mouth (11.8%), alveolar ridge (9.9%), retromandibular trigone (8.7%), buccal mucosa (7.5%), hard palate (1.9%), gingiva (1.2%), and lip mucosa (1.2%). In our cohort, 72.8% of patients (n= 118) were not actively taking ARB or ACEi pharmacotherapy. Among the others 20.4% were taking ACEis (n= 33), and 6.8% patients were taking ARBs (n= 11).

**Table 1.** Demographic characteristics and tumor related variables

| **Variable** |  | **ACEIs** | **ARBs** | **None** | **p-value** |
| --- | --- | --- | --- | --- | --- |
| **Age, Mean (SD)** |  | 63.09(11.95) | 66.9(7.93) | 57.35(14.87) | 0.014 |
| **Gender** | Female | 6(18.2) | 1(9.1) | 31(26.3) | 0.359 |
|  | Male | 27(81.8) | 10(90.9) | 87(73.7) |  |
| **Body mass index, n (%)** | ≤25 | 9(27.3) | 4(36.4) | 50(42.7) | 0.273 |
|  | >25 | 24(72.7) | 7(63.6) | 67(57.3) |  |
| **ASA, n (%)** | 2 | 2(6.1) | 0.(0) | 20(17.1) | 0.132 |
|  | 3/4 | 31(93.9) | 11(100) | 97(82.9) |  |
| **History of smoking, n (%)** | Current | 11(45.8) | 4(66.7) | 51(68) | 0.162 |
|  | Past history | 11(45.8) | 2(33.3) | 23(30.7) |  |
|  | None | 2(8.3) | 0.(0) | 1(1.3) |  |
| **Tumor location, n (%)** | Tongue | 13(39.4) | 3(27.3) | 75(64.1) | 0.006 |
|  | Other | 20(60.6) | 8(72.7) | 42(35.9) |  |
| **Tumor staging, n (%)** | 1/2 | 16(48.5) | 6(54.5) | 43(36.4) | 0.291 |
|  | 3/4 | 17(51.5) | 5(45.5) | 75(63.6) |  |
| **Node staging, n (%)** | No | 14(42.4) | 5(45.5) | 57(48.3) | 0.828 |
|  | N1-N4 | 19(57.6) | 6(54.5) | 61(51.7) |  |
| **Perineural invasion, n (%)** | No | 23(69.7) | 10(90.9) | 66(55.9) | 0.035 |
|  | Yes | 10(30.3) | 1(9.1) | 52(44.1) |  |
| **Neoadjuvant chemotherapy, n (%)** | No | 28 (84.8) | 10(90.9) | 92(98) | 0.445 |
|  | Yes | 5(15.2) | 1(9.1) | 26(22) |  |
| **Any analgesic, n (%)** | No | 15 (45.5) | 6 (54.5) | 46 (39) | 0.542 |
|  | Yes | 18 (54.5) | 5 (45.5) | 72 (61) |  |
| **NSAIDs, n (%)** | No | 31 (93.9) | 10 (90.9) | 105 (89.9) | 0.693 |
|  | Yes | 2 (6.1) | 1 (9.1) | 13 (11.1) |  |

ACEIs: Angiotensin converting enzyme inhibitors; ARBs: Angiotensin receptor blockers. ASA: American Society of Anesthesiologists. NSAIDs: Non-steroidal anti-inflammatory drugs.

From Kruskal-Wallis test or Fisher’s exact test, we observed statistically significant differences in age, tumor location and perineural invasion among treatment groups (Table 1). Patients taking ARBs had the highest median age when compared to the other two groups (65 vs. 62 for ACEi and 58 for none; p= 0.015). Interestingly, patients taking ARBs also had the lowest rate of perineural invasion (9.1% vs. 30.3% for ACEi and 44.1% for none; p = 0.035), as well as the lowest percentage of tumoral location at the tongue (27.3% vs 39.4% for ACEi and 64.1% for none; p = 0.006). We did not detect a significant difference in rate of neoadjuvant chemotherapy, analgesics, or NSAID use among different treatment groups.

### Oral cancer pain by pharmacotherapies (ACEis, ARBs, or none)

In our cohort, the majority of patients (73.1%, n = 117) experienced mild levels of preoperative pain (VNRS: 0-3), whereas 16.3% of patients (n= 26) reported moderate levels of preoperative pain (VNRS: 4-6), and only 10.6% (n= 17) reported severe pain (VNRS ≥7). The rate of moderate to severe preoperative pain was higher for patients with stage N1-N4 tumors compared to patients presenting in stage N0 (34.1% vs 18.7%; p = 0.028). Furthermore, our univariate analysis indicated that individuals with tumors with perineural invasion more commonly reported severe pain (p = 0.017). Patients taking ARBs were found to have the lowest level of perineural invasion (90.0% vs 69.7% in ACEi and 55.9% in none, p = 0.351) (Table 1).

Our analysis did not demonstrate a statistically significant change in pain intensity when comparing the use of ARBs or ACEis to the no treatment group (p =0.251) (Table 2). Due to the small sample of patients reporting severe pain (VNRS ≥7; n = 17), our multivariable logistic regression model was limited to two covariates. With the adjustment of tumor location, the odds of severe preoperative pain for patients with perineural invasion was 3.938 times higher (OR=3.938, 95% CI for OR: 1.335, 11.615; p-value=0.013) than patients without perineural invasion (Table 3).

**Table 2.** Effect of ACEIs and ARBs on preoperative pain and inflammatory scores

| **Variable** | **ACEIs**<br>Mean (SD) | **ARBs**<br>Mean (SD) | **None**<br>Mean (SD) | **p-value** |
| --- | --- | --- | --- | --- |
| **Pain VNRS** | 1.66 (2.94) | 2.72 (2.61) | 1.91 (2.71) | 0.251 |
| **NLR** | 2.79 (1.61) | 3.18 (2.24) | 2.94 (2.58) | 0.701 |
| **MLR** | 0.4 (0.19) | 0.52 (0.67) | 0.39 (0.22) | 0.869 |

VNRS: verbal numeric rating scale. NLR: neutrophil-to-lymphocyte ratio. MLR: Monocyte-to-lymphocyte ratio. ACEIs: Angiotensin converting enzyme inhibitors. ARBs: Angiotensin receptor blockers

**Table 3.** Multivariable analysis of factors associated with severe pain

| **Odds Ratio Estimates and Wald Confidence Intervals** |  |  |  |  |
| --- | --- | --- | --- | --- |
| **Effect** | **p-value** | **OR** | **95% CI for OR** |  |
| **Tumor location: Other vs. Tongue** | 0.0364 | 3.182 | 1.076 | 9.414 |
| **Perineural Invasion: Yes vs. No** | 0.0130 | 3.938 | 1.335 | 11.615 |

OR: Odds ratio. CI: Confidence Interval

### Inflammatory biomarkers (NLR and MLR) by pharmacotherapies (ACEis, ARBs, or none)

In analyzing the effects of ARBs and ACEis on the inflammatory pathway, we utilized biomarkers, NLR and MLR, to indicate levels of leukocyte proliferation. The median of preoperative NLR and MLR were 2.32 and 0.35, respectively, which are within normal clinical limits. As shown in Table 2, Kruskal-Wallis test did not indicate a significant difference in levels of preoperative biomarkers NLR (p = 0.701) or MLR (p = 0.869) among treatment groups. Likewise, the preoperative NLR (p=0.231) and MLR (p=0.933) were not found to be significantly different among patients with severe preoperative pain when compared to patients with mild to moderate preoperative pain (Table 2). Interestingly, patients taking ARBs had the highest mean preoperative NLR (3.187) and preoperative MLR (0.529) when compared to the ACEi and no treatment group (Table 2). Although these differences were not statistically significant, contrary to our hypothesis, patients taking ARBs presented with slightly higher levels of inflammatory biomarkers.

## Discussion

Pain remains an unresolved health problem in patients with oral cancer. Our study suggests that ACEis and ARBs are not associated with significant analgesic effects when compared no treatment. The association between the use of ARBs and functional outcomes has been investigated in other human pain conditions including total knee arthroplasty, migraine and chemotherapy-induced neuropathy.[16–18] Patients using ARBs did not show any improvement in knee flexion scores after total knee arthroplasty and in volunteers with experimental ischemic pain.[4, 16] However, in other pathologies like migraine, Diener et al. demonstrated that telmisartan reduced pain severity compared to placebo.[17] Interestingly, the use of ACEis and ARBs was associated with some protection of myelinated fiber function in a mixed population of patients with chemotherapy-induced neuropathy.[18]

There are several potential explanations for our findings. First, Scheff et al. reported that inflammation is a main driver of the mechanism of pain in oral cancers.[10] In support of that notion, a retrospective study demonstrated non-steroidal anti-inflammatory drugs showed some analgesic effects in patients with oral cancer.[19] The modulatory effects of ARBs on the inflammatory response are well documented; however their anti-inflammatory activity appears to be drug specific (telmisartan > ibesartan > valsartan and losartan).[20, 21] In our study, we investigated whether ARBs or ACEis had any impact on inflammatory scores including NLR and MLR. Interestingly, we found no difference in those markers of inflammation. We can speculate that our results are confounded by the fact that we grouped several ARBs in the analysis; thus, we were not able to detect a significant effect on inflammatory scores.

Secondly, the selectivity of ARBs against the angiotensin II type I receptors (AT2R1) and angiotensin II type II receptors (AT2R2) should be considered. With the exception of losartan, all ARBs are highly selective for the AT2R1 and show 10,000–30,000 times greater affinity for this receptor than for AT2R2. As a result of ARBs high selectivity, the AT2R2 can be exposed to a higher concentration of ATII counteracting the effects of AT2R1.[22] Additionally, previous preclinical studies have indicated that the proinflammatory effects of the renin-angiotensin system result from AT2R1 stimulation at the target organ, and thus, it is plausible that systemic blockade of AT2R1 signaling has opposing effects on different tissues.[23]

Thirdly, patients in our cohort taking ARBs were less likely to have perineural invasion, and tumors without perineural invasion were less likely to report severe pain. However, we did not find a significant reduction in severe pain among patients taking ARBs. It is important to consider that perineural invasion is a complex process highly regulated by multiple nociceptive mediators which are predominantly inflammatory cytokines, chemokines, and growth factors.[24] Although perineural invasion encompasses both inflammatory and neuropathic insults, oral cancers ARBs appear to be more effective in the modulation of neuropathic than inflammatory pain which may explain the lack of significant effect in pain improvement.[22] Shepherd et al. demonstrated that ARBs ameliorated pain behaviors in animals with spared nerve injury rather than those with inflammatory pain.[15] In that study, the authors suggested that angiotensin II receptors (AT2R) signaling on peripheral macrophages was an indispensable drive for the development of chronic neuropathic pain but not inflammatory pain.[15] Other investigations support Shepherd’s findings. For instance, ARBs showed anti-allodynic effects in animals with vincristine and paclitaxel induced neuropathic pain.[7, 25] In our cohort, the level of bioinflammatory markers, NLR and MLR, did not differ significantly among ACEi and ARB treatment groups when compared to no treatment, again supporting the lack of efficacy of ARBs on inflammatory pain.

Lastly, previous data suggests that angiotensin converting enzyme inhibitors inhibit the enzyme dipeptidyl carboxypeptidase thus, blocking the degradation of nociceptive kinins such as bradykinin and substance P.[26, 27] In animals with paclitaxel-induced neuropathy, enalapril worsened neuropathic pain by increasing the concentrations of bradykinin-related peptides in the sciatic nerve.[28] Contrarily, ramipril decreased the pain perception threshold to ischemic pain in humans and hypertensive rats as well as in those with neuropathic pain after constriction nerve injury; suggesting a potential analgesic effect of ACEis.[4, 5, 29] However, these medications did not have any impact on acute postsurgical pain in a large cohort of patients.[30] Similarly, our study suggest that the use of ACEis does not have an important effect in the expression of pain.

Our work has several limitations including the retrospective design and the small number of patients receiving ARBs and ACEis which may have affect the statistical analysis. Furthermore, due to the small number of patients receiving those medications, dose-response studies or any investigation to underpin their effect on our primary outcome was not possible. Another limitation of this study is that we did not account for differences in baseline levels of plasma AT2 between treatment and control groups. AT2 is known to be elevated in patients with hypertension.[31] However, the patients in our control group consisted of both hypertensive and normotensive patients. Thus, it may be plausible that the elevated AT2 levels in hypertensive patients taking ACEis and ARBs were simply reduced to normotensive levels, negating any differences in potential anti-inflammatory or analgesic effects between treatment and control groups.

In conclusion, our study suggests that the use of ACEis or ARBs is not associated with a significant effect on the expression of pain in patients with oral cancers. However, patients taking ARBs were shown to have lower levels of perineural invasion. Additional studies should be performed to establish whether ARB-induced reduction in perineural invasion is due to direct effect on cancer cells or via an indirect effect on infiltration leukocytes.

## Data Availability

All data referred to in the manuscript is securely stored and available to authors and co-authors of this work. This work is only in pre-preprint status.

## Conflict of interest

The author(s) declare that they have no conflict of interest.

## Notes

### Competing Interest Statement

The authors have declared no competing interest.

## References

1. Bray F, Ferlay J, Soerjomataram I, Siegel RL, Torre LA, Jemal A. Global cancer statistics 2018: GLOBOCAN estimates of incidence and mortality worldwide for 36 cancers in 185 countries. CA Cancer J Clin. 2018;68(6):394–424.

2. Elrefaey S, Massaro MA, Chiocca S, Chiesa F, Ansarin M. HPV in oropharyngeal cancer: the basics to know in clinical practice. Acta Otorhinolaryngol Ital. 2014;34(5):299–309.

3. Salvador GLO, Marmentini VM, Cosmo WR, Junior EL. Angiotensin-converting enzyme inhibitors reduce mortality compared to angiotensin receptor blockers: Systematic review and meta-analysis. European Journal of Preventive Cardiology. 2017;24(18):1914–24.

4. Kalra J, Chaturvedi A, Kalra S, Chaturvedi H, Dhasmana DC. Modulation of pain perception by ramipril and losartan in human volunteers. Indian J Physiol Pharmacol. 2008;52(1):91–6.

5. Kaur P, Muthuraman A, Kaur J. Ameliorative potential of angiotensin-converting enzyme inhibitor (ramipril) on chronic constriction injury of sciatic nerve induced neuropathic pain in mice. Journal of the Renin-Angiotensin-Aldosterone System. 2014;16(1):103–12.

6. Khan N, Muralidharan A, Smith MT. Attenuation of the Infiltration of Angiotensin II Expressing CD3+ T-Cells and the Modulation of Nerve Growth Factor in Lumbar Dorsal Root Ganglia—A Possible Mechanism Underpinning Analgesia Produced by EMA300, An Angiotensin II Type 2 (AT2) Receptor Antagonist. Frontiers in Molecular Neuroscience. 2017;10.

7. Kim E, Hwang S-H, Kim H-K, Abdi S, Kim HK. Losartan, an Angiotensin II Type 1 Receptor Antagonist, Alleviates Mechanical Hyperalgesia in a Rat Model of Chemotherapy-Induced Neuropathic Pain by Inhibiting Inflammatory Cytokines in the Dorsal Root Ganglia. Molecular Neurobiology. 2019;56(11):7408–19.

8. List MA, Stracks J, Colangelo L, Butler P, Ganzenko N, Lundy D, et al. How Do Head and Neck Cancer Patients Prioritize Treatment Outcomes Before Initiating Treatment? Journal of Clinical Oncology. 2000;18(4):877-.

9. Dios PD, Leston JS. Oral cancer pain. Oral oncology. 2010;46(6):448–51.

10. Scheff NN, Ye Y, Bhattacharya A, MacRae J, Hickman DN, Sharma AK, et al. Tumor necrosis factor alpha secreted from oral squamous cell carcinoma contributes to cancer pain and associated inflammation. Pain. 2017;158(12):2396–409.

11. Ho A, Wong C, Lam C. Tumor necrosis factor-α up-regulates the expression of CCL2 and adhesion molecules of human proximal tubular epithelial cells through MAPK signaling pathways. Immunobiology. 2008;213(7):533–44.

12. Bakst RL, Xiong H, Chen C-H, Deborde S, Lyubchik A, Zhou Y, et al. Inflammatory Monocytes Promote Perineural Invasion via CCL2-Mediated Recruitment and Cathepsin B Expression. Cancer Res. 2017;77(22):6400–14.

13. Ye Y, Dang D, Zhang J, Viet CT, Lam DK, Dolan JC, et al. Nerve Growth Factor Links Oral Cancer Progression, Pain, and Cachexia. Molecular cancer therapeutics. 2011;10(9):1667–76.

14. Anand U, Yiangou Y, Sinisi M, Fox M, MacQuillan A, Quick T, et al. Mechanisms Underlying Clinical Efficacy of Angiotensin II Type 2 Receptor (AT2R) Antagonist EMA401 in Neuropathic Pain: Clinical Tissue and in Vitro Studies. Molecular Pain. 2015;11.

15. Shepherd AJ, Mickle AD, Golden JP, Mack MR, Halabi CM, de Kloet AD, et al. Macrophage angiotensin II type 2 receptor triggers neuropathic pain. Proceedings of the National Academy of Sciences. 2018;115(34):E8057-E66.

16. Langston JR, Ramsey DC, Skoglund K, Schabel K. Angiotensin II blockade had no effect on range of motion after total knee arthroplasty: a retrospective review. Journal of Orthopaedic Surgery and Research. 2020;15(1).

17. Diener HC, Gendolla A, Fruersenger A, Evers S, Straube A, Schumacher H, et al. Telmisartan in Migraine Prophylaxis: A Randomized, Placebo-Controlled Trial. Cephalalgia: an international journal of headache. 2009;29(9):921–7.

18. Roldan CJ, Song J, Engle MP, Dougherty PM. Angiotensin-Converting Enzyme Inhibitors and Angiotensin Receptor Blockers Modulate the Function of Myelinated Fibers after Chemotherapy: A Quantitative Sensory Testing Study. Pain Physician. 2017;20(4):281–92.

19. Derry S, Wiffen PJ, Moore RA, McNicol ED, Bell RF, Carr DB, et al. Oral nonsteroidal anti-inflammatory drugs (NSAIDs) for cancer pain in adults. Cochrane Database Syst Rev. 2017.

20. Arjmand M-H, Zahedi-Avval F, Barneh F, Mousavi SH, Asgharzadeh F, Hashemzehi M, et al. Intraperitoneal Administration of Telmisartan Prevents Postsurgical Adhesion Band Formation. Journal of Surgical Research. 2020;248:171–81.

21. Song K-H, Park J-H, Jo I, Park J-Y, Seo J, Kim SA, et al. Telmisartan attenuates hyperglycemia-exacerbated VCAM-1 expression and monocytes adhesion in TNFα-stimulated endothelial cells by inhibiting IKKβ expression. Vascular Pharmacology. 2016;78:43–52.

22. Miura S-i, Karnik SS, Saku K. Review: Angiotensin II type 1 receptor blockers: class effects versus molecular effects. Journal of the Renin-Angiotensin-Aldosterone System. 2010;12(1):1–7.

23. Crowley SD, Rudemiller NP. Immunologic Effects of the Renin-Angiotensin System. Journal of the American Society of Nephrology. 2017;28(5):1350–61.

24. Amit M, Na’ara S, Gil Z. Mechanisms of cancer dissemination along nerves. Nat Rev Cancer. 2016;16(6):399–408.

25. Bessaguet F, Danigo A, Bouchenaki H, Duchesne M, Magy L, Richard L, et al. Neuroprotective effect of angiotensin II type 2 receptor stimulation in vincristine-induced mechanical allodynia. Pain. 2018;159(12):2538–46.

26. Rohit, Rao C, Krishna G. Effects of captopril and losartan on thermal and chemical induced pain in mice. Indian J Physiol Pharmacol. 2006;50(2):169–74.

27. Silva CR, Oliveira SM, Hoffmeister C, Funck V, Guerra GP, Trevisan G, et al. The role of kinin B1receptor and the effect of angiotensin I-converting enzyme inhibition on acute gout attacks in rodents. Annals of the Rheumatic Diseases. 2016;75(1):260–8.

28. Brusco I, Silva CR, Trevisan G, de Campos Velho Gewehr C, Rigo FK, La Rocca Tamiozzo L, et al. Potentiation of Paclitaxel-Induced Pain Syndrome in Mice by Angiotensin I Converting Enzyme Inhibition and Involvement of Kinins. Molecular Neurobiology. 2016;54(10):7824–37.

29. Aykan DA, Koca TT, Yaman S, Eser N. Angiotensin converting enzyme and neprilysin inhibition alter pain response in dexhamethasone-induced hypertensive rats. Pharmacological Reports. 2019;71(2):306–10.

30. Turan A, Atim A, Dalton JE, Keeyapaj W, Chu W, Bernstein E, et al. Preoperative Angiotensin-converting Enzyme Inhibitor Use is Not Associated With Increased Postoperative Pain and Opioid Use. Clin J Pain. 2013;29(12):1050–6.

31. Catt KJ, Cain MD, Zimmet PZ, Cran E. Blood Angiotensin II Levels of Normal and Hypertensive Subjects. Bmj. 1969;1(5647):819–21.

